# Effect of COVID-19 on Lipid Profile and its Correlation with Acute Phase Reactants

**DOI:** 10.1101/2021.04.13.21255142

**Authors:** Jahanzeb Malik, Talha Laique, Uzma Ishaq, Amna Ashraf, Asmara Malik, Mommana Ali, Syed Muhammad Jawad Zaidi, Muhammad Javaid, Asad Mehmood

## Abstract

**Background and Objective:** Coronavirus disease 2019 (COVID-19) manifests as multiple clinical and pathological organ dysfunctions. It also disrupts metabolic profile due to the release of pro-inflammatory cytokines causing a systemic inflammation reaction. However, the development and correlation of dyslipidemia with acute phase reactants is unknown. This investigation was performed to assess the pathological alterations of low-density lipoprotein cholesterol (LDL-C), high-density lipoprotein (HDL), triglycerides, and total cholesterol levels in COVID-19 patients.

**Methods:** This was a prospective study performed on real-world patients to assess serum levels of LDL-C, HDL, TG, TC on COVID-19 patients (mild: 319; moderate: 391; critical: 357) hospitalized at our center between April 2020 through January 2021. Age- and gender-matched controls who had their lipid profiles in the same period were included as the control group.

**Results:** LDL-C, HDL, TG, and TC levels were significantly lower in COVID-19 patients when compared with the control group (P < 0.001, 0.047, 0.045, < 0.001, respectively). All parameters decreased gradually with COVID-19 disease severity (LDL-C: median (IQR), mild: 98 (91,134); moderate: 97 (81,113); critical: 68 (68,83); HDL: mild: 45 (37,50); moderate: 46 (41,50); critical: 40 (37,46); TG: mild: 186 (150,245); moderate: 156 (109,198); critical: 111 (98,154); TC: mild: 224 (212,238); moderate: 212 (203,213); critical: 154 (125,187)). LDL-C, TC, and TG were inversely correlated with acute phase reactants (interleukin-6 (IL-6), Procalcitonin, C-reactive protein (CRP), and D-dimers). Logistic regression demonstrated lipid profile, thyroid profile, and acute phase reactants as predictors of severity of COVID-19 disease.

**Conclusion:** Hypolipidemia develops in increasing frequency with severe COVID-19 disease. It inversely correlates with levels of acute-phase reactants, indicating SARS-COV-2 as the causative agent for alteration in lipid and thyroid levels.

## Introduction

Coronaviruses include a large family of viruses notorious for causing a multitude of diseases in humans, ranging from common flu to more serious conditions like Middle Eastern Respiratory Syndrome (MERS) or Severe Acute Respiratory Syndrome (SARS) [1]. The causative agent for coronavirus disease 2019 (COVID-19) is the SARS coronavirus 2 (SARS-COV-2) that causes various manifestations of the disease, involving the respiratory, gastrointestinal, hematological, endocrine, and cardiovascular systems [2,3]. Even after the mitigation measure, the estimated basic reproduction (R0) ranges from 2.24 to 3.58 and the mortality rate of COVID-19 is considered to be 2.3% [4,5,6]. World Health Organization (WHO) declared COVID-19 as a pandemic and initiated a global emergency from 11th March 2020 [7].

SARS-COV-2 is a positive-stranded RNA virus with an envelope of lipid bilayer, and a genome of approximately 30,000 nucleotides. On an electron microscope four major proteins have been identified as an integral part of the virus: the nucleocapsids (N) protein, the crown or spike (S) protein, the membrane (M) protein, and the envelope (E) protein [8]. These proteins play an important role in human infectivity and among them, the S protein mediates attachment to human hosts via angiotensin-converting enzyme 2 (ACE2) receptors [9]. An expression of ACE 2, in combination with transmembrane protease serine 2 (TMPRESS2), seems to be the key cellular mechanism for human infection [10].

In SARS-COV-2, lipids are an essential component for maintaining the integrity of the viral membrane and its fusion to the host cells, viral replication, endo/exocytosis, and human host infiltration. Some preliminary reports have described lipid abnormalities associated with the severity of disease in COVID-19 [11,12]. Therefore, this investigation was aimed to assess the lipid profile and its correlation with other biochemical tests used to predict the severity of COVID-19.

## Methods

### Study design and patient demographics

This prospective study was carried out at Infectious Disease Block, Advanced Diagnostics and Liver Center, Rawalpindi, and was approved by the Review Committee (Study ID#ADC/20/006) of Advanced Diagnostics and Liver Center according to the ethical principles of the Declaration of Helsinki. Written, informed consent was taken from all the participants/guardians before data collection. A total of 1,755 patients were enrolled in this investigation: 1067 with COVID-19 and 688 healthy adults as controls from April 2020 through January 2021. All the data regarding demographics, epidemiology, comorbidities, laboratory tests, and hospital stay were extracted electronically. SARS-COV-2 positive patients were confirmed using real-time reverse transcription-polymerase chain reaction (RT-PCR). Pneumonia was classified according to the guidelines issued by the Centers for Disease Control (CDC) as labeled as mild, moderate, or severe on high resolution computed tomography (HRCT) scans according to our previous study [13]. HRCT was considered positive if it demonstrated consolidation, septal thickening, linear opacities, crazy-paving pattern, or halo sign. Mild disease was classified as less than 10% lung involvement with normal room oxygen saturation. Moderate cases were classified as less than 50% lung involvement on HRCT with respiratory rate of >30 breaths/min, and oxygen saturation of < 90% at rest requiring <10 ml/min of oxygen. Those requiring >10 ml/min of oxygen and/or ventilatory support were labeled as critical cases. Age- and gender-matched healthy adults, who had their lipid profile panel done at our clinic were included as controls. Their data were not identified and only age, gender, BMI, and lipid profile were extracted.

### Laboratory tests

All the laboratory tests were carries out at our certified laboratory in Advance Diagnostics under the standard procedures of the Punjab Healthcare Commission. Hemoglobin (Hgb), and white blood cells (WBC) were performed on a Sysmex automated hematology analyzer (XN-3100™). Alanine aminotransferase (ALT), aspartate aminotransferase (AST), low-density lipoprotein cholesterol (LDL-C), high-density lipoprotein (HDL), total cholesterol (TC), and triglycerides (TG) were tested on Cobas® c3011 analyzer (Roche Diagnostics). Free triiodothyronine (T3), free thyroxine (T4), thyroid-stimulating hormone (TSH), interleukin-6 (IL-6), and Procalcitonin were analyzed via electrochemiluminescent immunoassay (ECLIA) in the Elecsys® 2010 immunoassay system. All parameters were compared between the COVID-19 classification. In addition, the lipid profile was compared between the control group and COVID-19. All tests were drawn in a fasting state from the blood samples on admission.

### Statistical analysis

Data were analyzed with Statistical Package for the Social Sciences (SPSS) version 26 (IBM Corp, Armonk, NY, USA.). Normally distributed continuous variables were presented as mean ± standard deviation (SD) while Non-normally distributed variables as median (interquartile range). Categorical variables were presented in frequency and percentages. Normality test adjustment was applied through the Shapiro-Wilk test. The groups were compared using Student’s t-test (normal distribution) and the Mann-Whitney U test (non-normal distribution). Chi-square was used to compare categorical variables and the Kruskal-Wallis test was used to compare variables among multiple groups. A receiver operating characteristic (ROC) curve was calculated for all laboratory parameters to demonstrate sensitivity and specificity in the COVID-19 cohort. Odds ratio (OR) and confidence interval (CI) were presented for all tests performed. Scatter plots were demonstrated using Pearson correlation analysis. A p-value of less than 0.05 was considered significant.

## Results

A total of 1067 COVID-19 patients were included in this investigation: 319 were mild, 391 moderate, and 357 as critical cases. The ages for the COVID-19 cohort was approximately similar: mild (47.83 ± 19.66 years), moderate (47.87 ± 19.21 years), and critical (47.29 ± 18.76 years) while the mean age for age- and gender-matched control subjects was 57.37 ± 4.62 years. The patients in this group were significantly older than the COVID-19 cohort. Both COVID-19 and control groups had a majority of the male population (66.7% and 68.4%, respectively). **Table 1** demonstrates patient demographics and lab parameters in the control as well as COVID-19 groups. Forty-seven patients did not survive with a mortality rate of 4.4%. Biochemistry and hematology results showed that Hgb was significantly more in mild cases while WBC was increased in moderate and critical cases. AST, ALT, creatinine, CRP, D-dimers were all significantly increased in higher COVID-19 class while free T3, free T4, and TSH were markedly reduced in critical cases (**Table 1**). LDL-C, HDL, TC, and TG levels were significantly lower in COVID-19 patients as compared to control subjects. All lipid profiles decreased gradually and significantly in critical cases as compared with mild and moderate cases. **Figure 1** and **Table 2** demonstrate the receiver operating characteristic (ROC) curve exhibiting the sensitivity and specificity of the lab parameters in predicting the progression of COVID-19 disease. Correlation of LDL-C, HDL, TC, and TG with acute phase reactants is shown in **Figure 2**.

**Figure 1.**
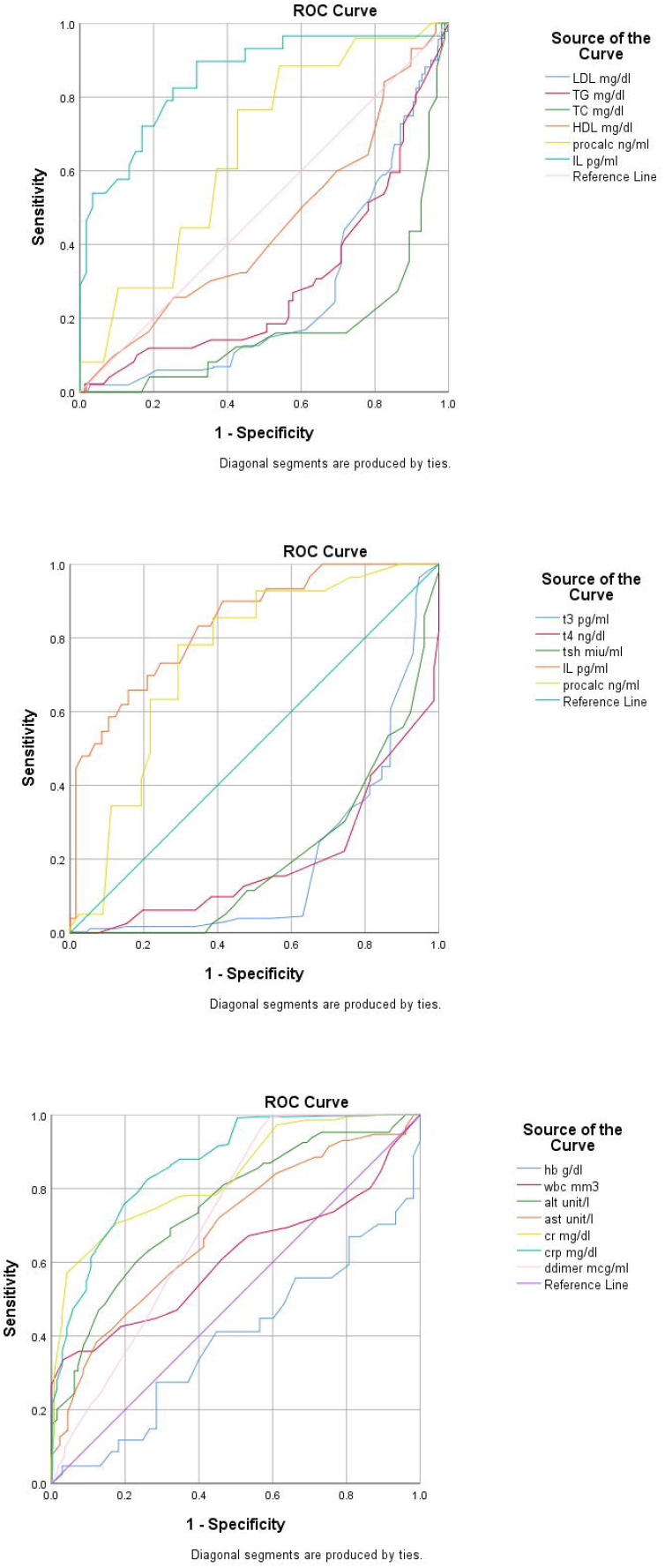
ROC curve analysis of lab parameters showing sensitivity and specificity in predicting severity of COVID-19. Receiver operating characteristic (ROC)

**Figure 2.**
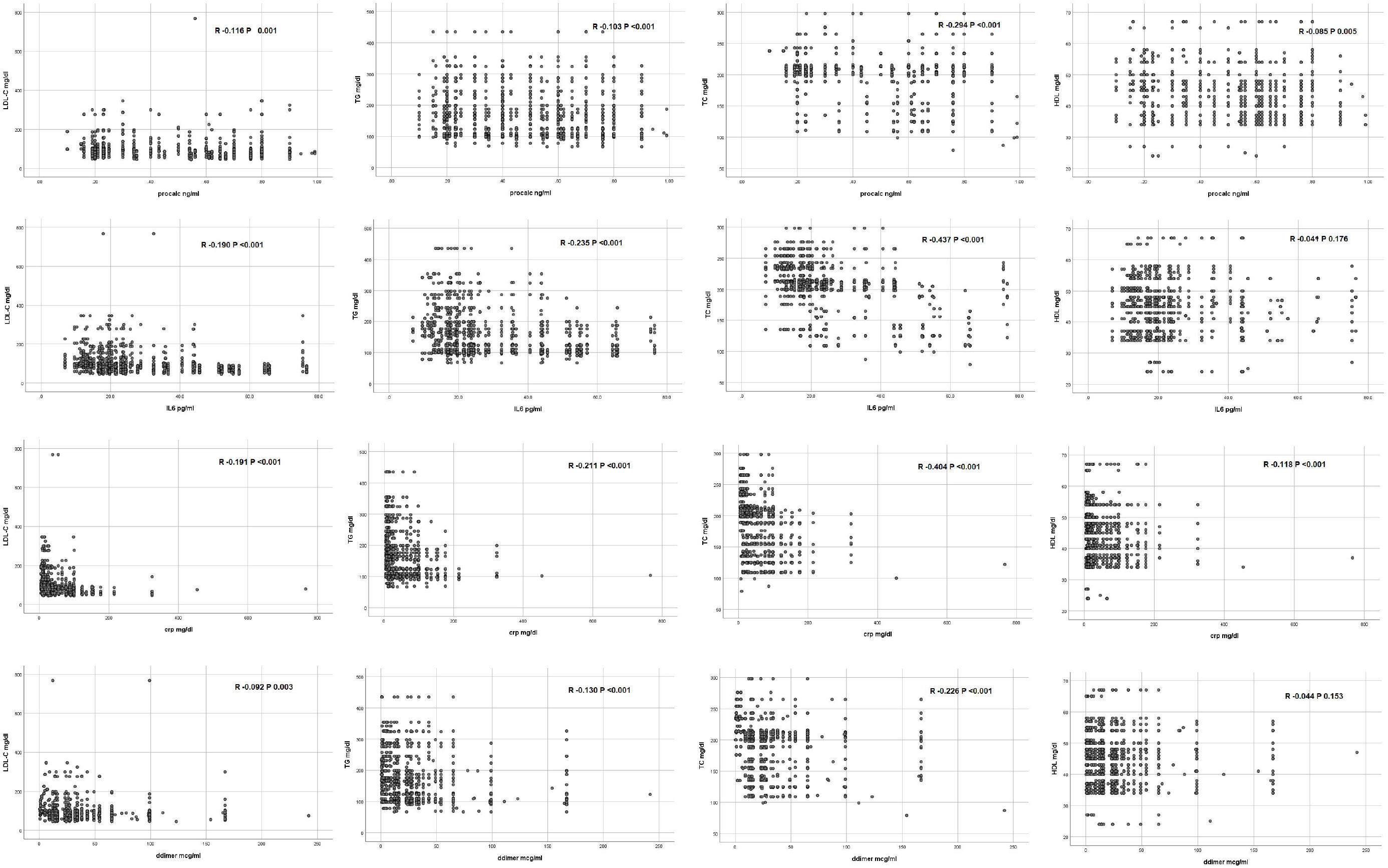
Correlation of lipid profile with acute phase reactants (interleukin-6, Procalcitonin, C-reactive protein, D-dimers)

Logistic regression analysis was performed to determine predictors of severe COVID-19 disease among all subjects. The model suggested LDL-C, HDL, TC, TG, free T3, free T4, TSH, IL-6, Procalcitonin, CRP, and D-dimers as independent risk factors for COVID-19 (**Table 3**).

## Discussion

Key findings of this investigation were as follows: Levels of LDL-C, HDL, TC, and TG all decreased significantly with increasing severity of COVID-19 while acute phase reactants like IL-6, Procalcitonin, CRP, and D-dimers were increased in this cohort and thyroid function tests, lipid profile parameters, and acute phase reactants were predictors of severe disease of COVID-19. This is the first prospective report with lipid profile data of real-world patients suffering from COVID-19 and this investigation points out that lipid levels start decreasing even in mild cases while reduced lipid levels inversely correlate with acute phase reactants. Also, it provides evidence of altered biochemistry and pathological evolution of lipidology in COVID-19.

Viral infections cause alterations of lipid biomarkers in their hosts, leading to disrupted cholesterol rafts. This helps them in infiltrating the host defenses. In response to acute inflammatory conditions in COVID-19, termed as cytokine storm, there is dysfunction of HDL and LDL-C particles. In addition, a surge of pro-inflammatory cytokines can cause a consumption of albumin, ApoA1, HDL, TC, TG, LDL-C, along with decreased lymphocytes [14]. Therefore, according to our study, the depressed lipid parameters can be a predictor for the severity of COVID-19 disease [11]. Similarly, a study in China described the lipid profile and other clinical characteristics of COVID-19 [15]. The levels of LDL-C, HDL, and TC were significantly decreased in their patients, and this alteration was gender-specific. In contrast to our study, males more commonly presented with lowered lipid levels in their investigation. A significant decrease in HDL was seen only in critical cases of COVID-19 while a significant decrease of LDL-C, TC, and TG was observed in all COVID-19 groups (mild, severe, critical). However, in our study HDL, LDL-C, TG, and TC were significantly depressed only in the critical COVID-19 cohort. In contrast to these investigations, a preprint reports significantly increased levels of LDL-C compared to control groups and a positive relationship of COVID-19 severity with elevated lipid profile alterations [16].

Similar findings have been demonstrated in human immunodeficiency virus (HIV), where acquired immunodeficiency syndrome (AIDS) patients exhibit an elevated level of plasma triglycerides and decreased HDL and LDL-C [17]. Furthermore, HIV-infected individuals have the propensity to develop various other lipid abnormalities like hypocholesterolemia and decreased free fatty acids [18]. A study analyzed lipid metabolism in SARS-recovered patients at 12 years’ interval and demonstrated hypertriglyceridemia and an elevated very-low-density lipoprotein (VLDL) cholesterol [19].

These findings can be explained by the composition of HDL, which contains esterified cholesterol, Apo-lipoproteins, and triglycerides. These lipid fragments impart an important role in small-vessel vasodilation, and in the reduction of oxidation and free radical formation, apoptosis, thrombosis, infection, and inflammation [20]. In addition to its contribution as an anti-inflammatory lipoprotein, HDL downregulates inflammatory mediators by inactivating T-cells and macrophages [21]. In COVID-19, a surge in pro-inflammatory cytokines confer the presence of systemic inflammation. HDL can deactivate this inflammatory cascade by inhibiting the activation of monocytes and neutrophils while maintaining an antioxidant function, allowing for the removal of oxidized lipids and neutralizing oxidative factors. This in turn mitigates inflammatory response in the host cells [22]. During a cytokine storm in COVID-19, HDL and LDL-C are oxidized, leading to an upregulation of immune activation [23,24]. Based on the immunomodulatory mechanism of HDL, we can consider immune regulation in COVID-19 as the primary cause for decreased lipid levels in this investigation.

There were several limitations of this investigation. First, the time of symptom onset to the time of sample collection was variable in this cohort. Therefore, the analysis might represent heterogeneous results of the COVID-19 disease course. Second, many patients were already on statin treatment and the level of precise alteration in lipid profile during the disease course of the COVID-19 cohort could be biased towards depressed lipid levels in these patients. Third, the control group patients were healthy individuals with no severe or critical form of acute illness. This could have produced a sample selection bias, producing abnormal results in this investigation. Fourth, no follow-up data of the lipid profile and its alteration was collected for monitoring the dynamics of COVID-19. This will be needed for better characterization of this phenomenon in the COVID-19 cohort.

## Conclusion

The ongoing COVID-19 outbreak caused by SARS-COV-2 poses a great challenge to the human population with a pressing need to understand viral mechanisms and develop effective antiviral agents. As cholesterol is involved in many vital cellular processes in regulating the viral entry into the host cell, the results of this investigation demonstrate a correlation of the severity of COVID-19 disease with altered lipid profile parameters. Decreased levels of LDL-C, HDL, TC, and TG validate the mechanism of immune upregulation by HDL and its byproducts, causing a negative correlation between lipids and acute phase reactants released as pro-inflammatory cytokines in COVID-19.

## Data Availability

Upon request

## Acknowledgement

None

## Ethics statement/Informed consent statement

Advanced Diagnostics and Liver Center gave express permission after approval from the review board of Advanced Diagnostics and Liver Center to collect data within its facilities (ID# ADC/20/006) for this investigation. All participants gave written/informed consent before data collection.

## Ethics Committee

Ethical review board of Advanced Diagnostics and Liver Center, Rawalpindi, Pakistan.

## Author contribution

JM: concept, methodology, formal analysis, critical review and final draft; TL: first draft and critical review and final draft; UI: supervision, data curation, first draft; AA: methodology, first draft; AM: first draft, formal analysis; MAR: first draft, critical review and final draft; SMJZ: first draft, methodology, data curation; MJ: data curation, methodology; AM: data cutation, methodology, critical review and final draft

**Supporting Documents 1. All data is available in supporting document S1**.

