## Supplementary material for "Effect of COVID-19 on Lipid Profile and its Correlation with Acute Phase Reactants": table 1

| **Variables** | **Reference range** | **Control (688)** | **COVID-19 patients (1067)** | | | | |
| --- | --- | --- | --- | --- | --- | --- | --- |
|  | | | **Mild (319)** | **Moderate (391)** | **Critical (357)** | **OR (95% CI)** | **P-value** |
| **Age, mean ± SD** | - | 57.37 ± 4.62 | 47.83 ± 19.66 | 47.87 ± 19.21 | 47.29 ± 18.76 | - | 1 |
| **Gender n (%)** |  |  |  |  |  |  | 0.144 |
| Male | - | 471 (68.4%) | 226 (70.8%) | 250 (63.9%) | 236 (66.1%) | - |  |
| Female | - | 217 (31.5%) | 93 (29.2%) | 141 (36.1%) | 121 (33.9%) | - |  |
| **BMI (kg/m^2^) mean ± SD** | - | 27.21 **±** 3.62 | 27.53 ± 3.71 | 27.46 ± 3.61 | 27.45 ± 3.61 |  | 0.971 |
| **Comorbid conditions n (%)** | | | | | | | |
| DM | - | - | 93 (29.2%) | 120 (30.7%) | 97 (27.2%) | - | 0.570 |
| HTN | - | - | 111 (34.8%) | 129 (33%) | 121 (33.9%) | - | 0.880 |
| CVD | - | - | 96 (30.1%) | 93 (23.8%) | 93 (26.1%) | - | 0.162 |
| CKD | - | - | 34 (10.7%) | 31 (7.9%) | 31 (8.7%) | - | 0.435 |
| Liver disease | - | - | 29 (9.1%) | 50 (12.8%) | 59 (16.5%) | - | **0.016** |
| **Hospital stay, days, mean ±** **SD** | - | - | 4 ± 1.91 | 11.65 ± 2.78 | 31.02 ± 7.65 | 6.14 (15.13-16.59) | **<0.01** |
| **Lab parameters, median (IQR)** | | | | | | |  |
| LDL-C (mg/dl) |  | 115 (92,135) | 98 (91,134) | 97 (81,113) | 68 (68,83) | 2.43 (97.56-104.51) | **<0.01** |
| HDL (mg/dl) |  | 47 (36,52) | 45 (37,50) | 46 (41,50) | 40 (37,46) | 7.49 (43.95-44.93) | **<0.01** |
| TC (mg/dl) |  | 165 (98,254) | 224 (212,238) | 212 (203,213) | 154 (125,187) | 5.12 (193.89-198.95) | **<0.01** |
| TG (mg/dl) |  | 190 (126,235) | 186 (150,245) | 156 (109,198) | 111 (98,154) | 1.65 (161.21-169.96) | **<0.01** |
| IL-6 (pg/ml) |  | - | 14.30 (12.20,18.90) | 21.70 (17.60,28.70) | 43.80 (22.90,54.90) | 3.09 (26.76-28.72) | **<0.01** |
| Procalcitonin (ng/ml) |  | - | 0.40 (0.20,0.50) | 0.34 (0.23,0.70) | 11.80 (10.20,13.70) | 0.63 (0.46-0.49) | **<0.01** |
| Hgb (g/dl) |  | - | 13 (11.60,15.30) | 11.90 (11.20,13.30) | 11.80 (10.20,13.70) | 0.79 (12.55-12.84) | 0.162 |
| WBC (mm^3^) |  | - | 9000 (7000,11000) | 12000 (6000,15000) | 12000 (6000,19000) | 1.24 (10.90-11.64) | 0.074 |
| ALT (unit/l) |  | - | 23 (15,45) | 65 (45,89) | 89 (56,187) | 23.64 (138.16-177.28) | **<0.01** |
| AST (unit/l) |  | - | 34 (23,65) | 65 (43,89) | 76 (45,117) | 1.88 (94.37-152) | **<0.01** |
| Creatinine (mg/dl) |  | - | 0.90 (0.70,1.50) | 1.20 (0.90,1.60) | 2 (1.50,3.80) | 5.71 (1.60-1.75) | **<0.01** |
| Free T3 (pg/ml) |  | - | 2.5 (2,3) | 2.10 (1.60,2.70) | 1 (0.90,1.90) | 2.84 (1.88-2.01) | **<0.01** |
| Free T4 (ng/dl) |  | - | 1.50 (0.90,1.80) | 0.90 (0.80,1.60) | 0.60 (0.40,0.80) | 1.66 (1.13-1.24) | **<0.01** |
| TSH (miu/dl) |  | - | 2.60 (1.80,3.80) | 0.90 (0.80,1) | 0.80 (0.50,0.90) | 2.43 (1.44-1.63) | **<0.01** |
| CRP (mg/dl) |  | - | 9 (8,13) | 23 (11,29) | 65 (27,100) | 3.79 (37.32-43.79) | **<0.01** |
| D-dimers (mcg/ml) |  | - | 2 (1,5) | 24 (15,38) | 24 (15,40) | 1.15 (24.01-27.76) | **<0.01** |

**Table 1. Patient demographics and lab parameters. Normally distributed variables expressed as mean ± SD, abnormally distributed variables expressed as median (IQR). OR and 95%CI expressed between control and COVID-19 cohort. Categorical variables presented as n (%). P < 0.05 as significant. Odds ratio (OR), confidence interval (CI), standard deviation (SD), interquartile range (IQR), body mass index (BMI), diabetes mellitus (DM), hypertension (HTN), cardiovascular disease (CVD), chronic kidney disease (CKD), low-density lipoprotein (LDL), high-density lipoprotein (HDL), total cholesterol (TC), triglycerides (TG), interleukin (IL), hemoglobin (Hgb), white blood count (WBC), alaninie aminotransferase (ALT), aspartate aminotransferase (AST), triiodothyronine (T3), thyroxine (T4), thyroid stimulating hormone (TSH), C-reactive protein (CRP)**
