## Supplementary material for "Effect of COVID-19 on Lipid Profile and its Correlation with Acute Phase Reactants": table 2

| **Variable** | **AUC** | **Sensitivity** | **Specificity** | **P-value** |
| --- | --- | --- | --- | --- |
| **LDL-C** | 0.266 | 0.524 | 0.787 | **<0.01** |
| **HDL** | 0.448 | 0.301 | 0.353 | **<0.01** |
| **TC** | 0.173 | 0.041 | 0.189 | **<0.01** |
| **TG** | 0.298 | 0.270 | 0.578 | **<0.01** |
| **Free T3** | 0.197 | 0.608 | 0.869 | **<0.01** |
| **Free T4** | 0.194 | 0.098 | 0.442 | **<0.01** |
| **TSH** | 0.205 | 0.597 | 0.923 | **<0.01** |
| **IL-6** | 0.855 | 0.323 | 0.017 | **<0.01** |
| **Procalcitonin** | 0.659 | 0.082 | 0.064 | **<0.01** |

**Table 2. AUC, sensitivity, and specificity of lab parameters in correlation with the ROC curve. Area under the curve (AUC), low-density lipoprotein (LDL), high-density lipoprotein (HDL), total cholesterol (TC), triglycerides (TG), interleukin (IL), triiodothyronine (T3), thyroxine (T4), thyroid stimulating hormone (TSH)**
