## Supplementary material for "Effect of COVID-19 on Lipid Profile and its Correlation with Acute Phase Reactants": table 3

| **Variable** | **B** | **SE** | **Wald** | **P-value** |
| --- | --- | --- | --- | --- |
| **LDL-C** | 0.009 | 0.002 | 14.474 | **<0.01** |
| **HDL** | 0.143 | 0.029 | 23.627 | **<0.01** |
| **TC** | 0.167 | 0.019 | 74.122 | **<0.01** |
| **TG** | 0.018 | 0.004 | 19.194 | **<0.01** |
| **Free T3** | 0.441 | 0.096 | 21.242 | **<0.01** |
| **Free T4** | 1.996 | 0.449 | 19.739 | **<0.01** |
| **TSH** | 4.017 | 0.567 | 50.183 | **<0.01** |
| **IL-6** | -0.067 | 0.008 | 67.426 | **<0.01** |
| **Procalcitonin** | -3.183 | 0.572 | 30.953 | **<0.01** |
| **CRP** | -0.035 | 0.004 | 81.200 | **<0.01** |
| **D-dimers** | -0.009 | 0.004 | 5.357 | **0.021** |

**Table 3. Logistic regression analysis showing predictors of critical COVID-19 disease. Low-density lipoprotein (LDL), high-density lipoprotein (HDL), total cholesterol (TC), triglycerides (TG), interleukin (IL), triiodothyronine (T3), thyroxine (T4), thyroid stimulating hormone (TSH), interleukin-6 (IL-6), C-reactive protein (CRP)**
